# Safeguarding Adolescent Mental Health in India (SAMA): study protocol for co-design and feasibility study of a school systems intervention targeting adolescent anxiety and depression in India

**DOI:** 10.1101/2021.06.25.21259538

**Authors:** S. Hugh-Jones, J. Naidu, H. Al-Janabi, P. Bhola, P. Cook, M. Fazel, K. Hudson, P. Khandeparkar, T. Mirzoev, S. Venkataraman, R. West, P. Mallikarjun

## Abstract

**Introduction:** Symptoms of anxiety and depression in Indian adolescents are common. Schools can be opportune sites for delivery of mental health interventions. India, however, is without a whole-school mental health approach. This article describes the study design for the Safeguarding Adolescent Mental Health in India (SAMA) project. The aim of SAMA is to co-design and feasibility test a suite of multi-component interventions across the intersecting systems of adolescents, schools, families and their local communities in India.

**Methods and analysis:** Our project will co-design and feasibility test four interventions to run in parallel in eight schools (three assigned to waitlist) in Bengaluru and Kolar in Karnataka, India. The primary aim is to reduce the prevalence of adolescent anxiety and depression. Co-design of interventions will build on existing evidence and resources. Interventions for adolescents at school will be universal, incorporating curriculum and social components. Interventions for parents and teachers will target mental health literacy, and also for teachers, training in positive behaviour practices. Intervention in the school community will target school climate to improve student mental health literacy and care. Intervention for the wider community will be via adolescent-led films and social media. We will generate intervention cost estimates, test outcome measures and identify pathways to increase policy action on the evidence.

**Ethics and dissemination:** Ethical approval has been granted by the National Institute of Mental Health Neurosciences Research Ethics Committee (NIMHANS/26^th^ IEC (Behv Sc Div / 2020/2021)) and the University of Leeds School of Psychology Research Ethics Committee (PSYC-221). Certain data will be available on a data sharing site. Findings will be disseminated via peer-reviewed journals and conferences.

**Article Summary:** *Strengths and Limitations:*

1. A key strength of this study is its focus on depression and anxiety prevention, both of which are the common adolescent mental health problems in India.
2. The study utilises a public mental health systems approach, which adopts a ‘person-in-context’ perspective and recognises multiple determinants of mental health.
3. Extensive co-design of interventions will promote cultural relevance and acceptability by key stakeholders, including adolescents, teachers, parents and school communities.
4. The Covid-19 pandemic makes this research more pertinent but may affect recruitment of the schools / adolescents or may lead to participation bias if the study processes are conducted online by excluding those with limited access or who lack technical skills.
5. The opt-in approach to consent may lead to participation bias.

## Introduction

Mental health conditions including depression and anxiety disorders are among the top ten causes of illness and disability in adolescents.[1] Half of all lifelong mental-health conditions have their onset in adolescence. [2] Poor mental health in adolescence is associated with poor physical health and lifelong disadvantage, especially if education is disrupted, impacting occupational and social trajectories.[3]

India has the largest adolescent population in the world (253 million).[4] An estimated 9.8 million Indian 13–17-year-olds (pooled prevalence of 7.3%) have a clinical mental-health condition [5] but levels seem considerably higher in urban areas and among school-going adolescents. [6] Although there is limited data on the treatment gap in adolescents, the overall treatment gap for mental disorders in India is around 90%. [5] The treatment gap and the effectiveness and cost-effectiveness of early intervention are driving investment into public mental health in efforts to develop scalable programmes that can reach adolescents before their symptoms progress to levels of a clinical disorder.[5] Tackling preventable and treatable health conditions, and improving the quality of education for adolescents, present the single best investments for health and wellbeing a low-and-middle income (LMIC) country can make. [7]

Schools have tremendous potential as platforms for public mental-health interventions, which can secure positive effects on mental health, including a reduction in anxiety and depression, and which can be delivered, sustained and scaled in LMICs. [7] School provision can increase access to interventions over and above existing mental healthcare systems, which in LMICs, are typically fragmented, under-resourced, and not tailored to adolescents. [8–10] Improving school experience may also promote longer participation in education, which reduces a number of risks for girls in particular. [8] The Lancet commission on adolescent wellbeing calls for investment in school mental health, bolstered by the rise in school attendance [8] including in India (97% enrolment). [11] India, however, is without an evidence-based, whole school mental health approach for adolescents.

Our project responds to this need by identifying existing school interventions which can be integrated, tailored and culturally adapted into a systems approach [12] in India through a process of co-production with stakeholders. Given the multiple determinants of adolescent mental health in India, [5] interventions will need to target individual and contextual factors. In India, a key contextual determinant of adolescent mental health is the nature of schooling, including the use of corporal punishment [13–15], extreme academic pressure [16,17], a lack of mental health literacy among adolescents, teachers and parents, [18–20] prevailing mental health stigma and bullying,[21] poorly supported staff, often with large class sizes and a lack of mental health support within schools.[22] Although the Government of India endorsed the WHO Health Promoting School model (2007) [23] progress on school mental health has been slow. To date, school initiatives in India have mostly targeted physical health and life skills. [24–27] There is an urgent need to accelerate improvements in mental health in Indian schools, from a safety and rights perspective [28], to reduce school risks to adolescent mental health and to improve population health. India has limited evidence-based, scalable school programmes, endorsed by policy, to support adolescent mental health. This is a critical care gap. [5,29] Our study aims to contribute to the evidence base about what works in Indian schools to support adolescent mental health.

There is some evidence for school-based universal mental interventions for depression and anxiety prevention, and mental health promotion, in High-Income Countries (HICs) [30,31]. The evidence in LMICs is promising though limited [8–10]. Three whole-school health and life-skills programmes with small mental health components have been tested in India. [24,26,32,33] One did not report any beneficial clinical outcomes [24] and one has yet to establish effectiveness. [32] The third is a health promotion programme targeting physical and sexual health, bullying, school climate, gender equality and depressive symptoms. [26]

Outcomes were positive, including for depressive symptoms, with benefits extended up to two years [33]. It did not, however, target anxiety nor include broad mental health promotion. This is a critical gap given the prevalence of anxiety disorders in adolescence and its co-morbidity with depression.

Our study **S**afeguarding **A**dolescent **M**ental he**A**lth in India (SAMA) will co-design, and feasibility test, a suite of school interventions in India targeting multiple risks and protective factors across the school system. The health objective of these interventions is to reduce symptoms of anxiety and depression in symptomatic adolescents and to promote the wellbeing all school-going adolescents. The SAMA study will take place between January 2021 and December 2023.

## Methods and analysis

### Study design

SAMA will run over 36 months and will deliver 8 work packages (WP) (see Figure 1).

**Figure 1:**
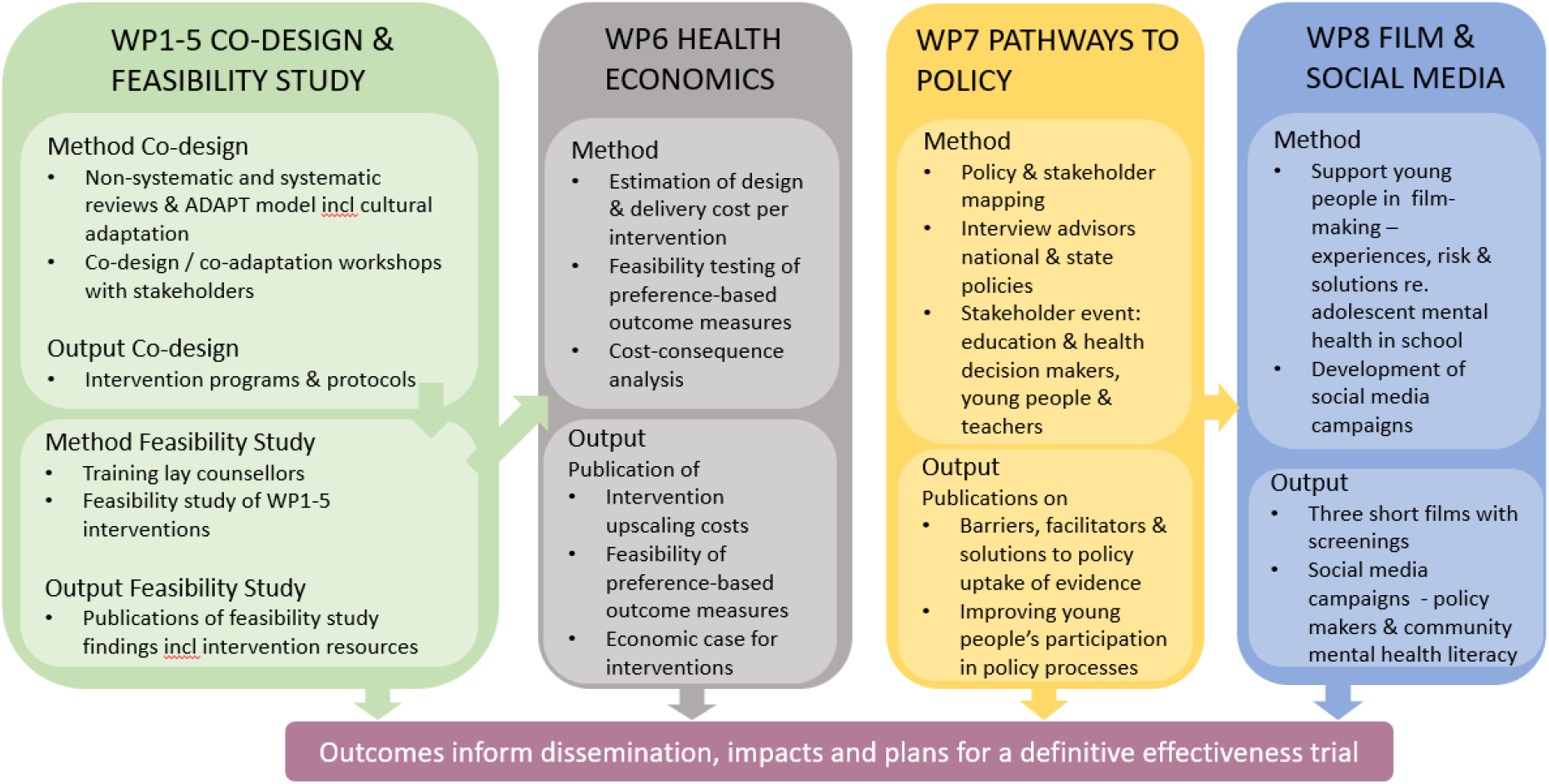
Study work packages.

**WP1-4 are intervention co-design / co-adaptation and feasibility studies following MRC guidance [34] and the Six Steps in Quality Intervention Development 6SQuID [35], targeting adolescents (14–15 years), teachers, the school community and parents (Table 1). WP5 is implementation research on these interventions and WP6 is an exploratory cost-effectiveness study of the interventions. WP7 and 8 target policy and local community systems respectively. Year 1 will focus on co-design and co-adaptation of interventions via WP1-5. Years 2 and 3 will include a feasibility study of the adapted interventions alongside WP6 and 7. WP8 will run across all three years**.

**Table 1:**
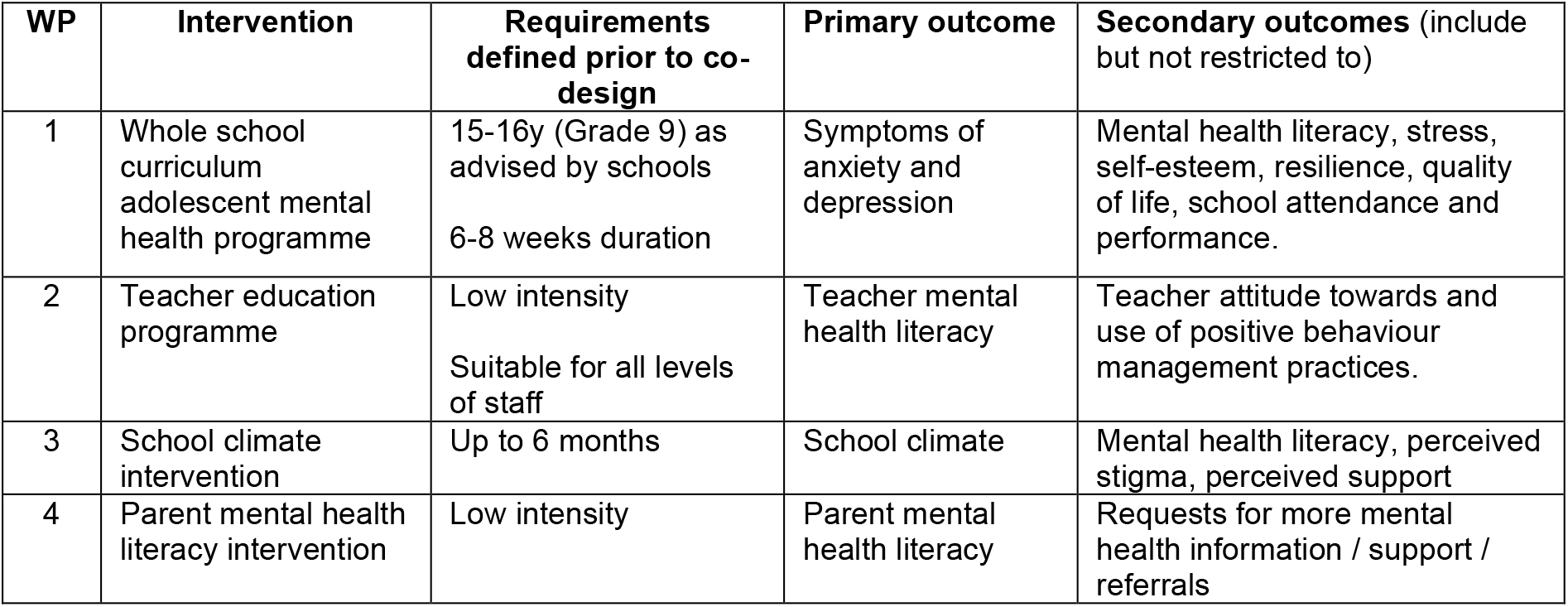
Intervention aims and requirements defined in advance.

| WP | Intervention | Requirements defined prior to co-design | Primary outcome | Secondary outcomes (include but not restricted to) |
| --- | --- | --- | --- | --- |
| 1 | Whole school curriculum adolescent mental health programme | 15-16y (Grade 9) as advised by schools<br>6-8 weeks duration | Symptoms of anxiety and depression | Mental health literacy, stress, self-esteem, resilience, quality of life, school attendance and performance. |
| 2 | Teacher education programme | Low intensity<br>Suitable for all levels of staff | Teacher mental health literacy | Teacher attitude towards and use of positive behaviour management practices. |
| 3 | School climate intervention | Up to 6 months | School climate | Mental health literacy, perceived stigma, perceived support |
| 4 | Parent mental health literacy intervention | Low intensity | Parent mental health literacy | Requests for more mental health information / support / referrals |

**SAMA will work with adolescents as partners in all stages of the project. In addition to the adolescents involved as participants in co-design, we will have a Youth Advisory Board who will be consulted on all project components, a subset of whom will form adolescent film crews and drive the social media campaigning (both WP8). We will have three other advisory boards (Steering Group, Scientific Advisory Board, Ethics Oversight Committee)**.

For WP1-4, some broad interventions requirements have been defined in advance of co-design, including the aims and primary and secondary outcomes. These are shown in Table 1. All interventions will be delivered by external lay counsellors. There is some evidence that this delivery approach is superior and preferred to teacher-delivered interventions, and peer educator programmes, at least for adolescent mental health programmes. [26] [22] [32] There are additional benefits of having a small team of lay counsellors working closely in partnership with schools.

### School recruitment

We will recruit eight schools in Kolar and Bangalore to participate in both the co-design and feasibility stages of SAMA (WP1–6). School recruitment will be facilitated by the Karnataka state primary and secondary education ministry and the Karnataka state secondary school Headmaster’s Association. We will recruit two school types (government and low-cost private) to identify school factors that may affect feasibility. Schools will be eligible to take part in SAMA if they endorse the delivery of WP1–6 in their schools, including participation in co-design. WP1 intervention feasibility testing will be with a new Grade 9 cohort not involved in co-design.

### Year 1 Co-design / co-adaptation of interventions

Co-design will proceed in stages (see Figure 2) and will include co-adaptation of existing interventions as well as design of new components where needed (e.g. content, implementation, safeguarding). We begin with co-adapting interventions before identifying the need for new co-designed elements. Our approach to co-adaptation is informed by phases 1–3 of the ADAPT model for introducing complex public health interventions into new settings [36] and also by Perera et al.’s approach [37] to adapting low-intensity psychological interventions into new cultural settings. WP5 runs alongside WP1–4 and aims to identify implementation facilitators and barriers for all intervention components. As well as adapting interventions, we will co-produce with adolescent and stakeholders a whole-school safeguarding protocol tailored to the interventions.

**Figure 2:**
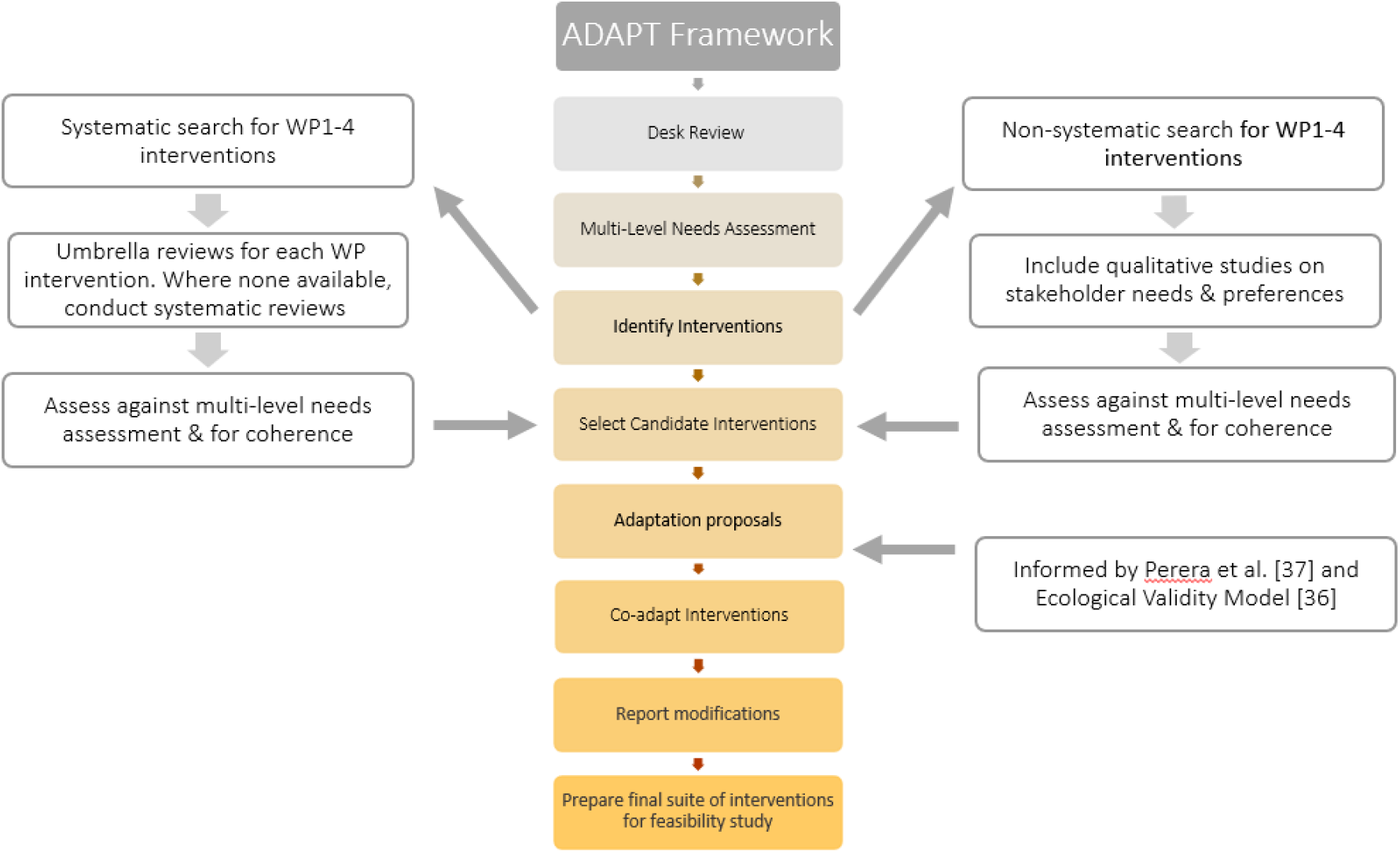
Stages of co-adaptation / co-design.

#### Phase 1: Exploration of needs and existing interventions

##### Initial assessment

A desk review will be conducted to gather knowledge on the local context and research evidence relevant to our project aims, including identification of any existing best practice and policy requirements. This will feed into an evidence-based, multi-level needs assessment of end-users, families, schools and regional / state systems to sensitise our selection of interventions and implementation approach.

##### Intervention selection & exploration

We aim to identify interventions that are likely to be acceptable, feasible and efficacious in our target context to meet our project aims. We will search for interventions with the potential (after co-adaptation) to impact at least the primary outcome for each WP (Table 1). As per Figure 2, a non-systematic search will be conducted first to collate or identify interventions which are known to the team in India, referenced in Indian education or mental health policies or referenced by Indian or global organisations (e.g. Save the Children, WHO). This will be followed by a systematic search. Umbrella review protocols for these are registered on Prospero [38–41] and reviews are nearing completion. Considering search returns from both the non-systematic and systematic searched, WP1–4 leads will propose candidate interventions to be taken to Phase 2. Whilst established effectiveness of an intervention is important, we also need to consider the coherence, integration, implementation and scalability of interventions for the local context. [42] Proposals will be reviewed by the research team and project advisory boards, which include in-country adolescents, schools and mental health professionals as well as the team which led SEHER [26]. Amendments to proposals will be made where necessary.

This phase includes work on implementation protocols (WP5). We will build on a UK framework for a whole-school mental health intervention [43] based on the Consolidated Framework for Implementation Research (CFIR) [44]. The framework will be iteratively developed as SAMA interventions are finalised, drawing on existing implementation evidence from India, including the SEHER study [26], and through our co-adaptation processes.

#### Phase 2: Preparation for co-design

##### Identification of mismatches and model development

Candidate interventions will be assessed against our multi-level needs assessment; those failing to meet significant need will be rejected. We will then develop a model of how the final interventions could work across systems before identifying the aspects (across surface and deep levels [45]) which are most critical to take forward to co-design / co-adaptation.

Decisions here will be informed by the approach of Perera et al. [37] and their use of the Ecological Validity Model [46]. This will lead to the generation of adaptation hypotheses, that is, our proposals for which intervention aspects will need to be culturally adapted. The Ecological Validity Model prompts directs consideration of the language, persons, metaphors, content, concepts, goals, methods and context (which includes implementation) of the intervention to be adapted. We will supplement these proposals with co-design needs, that is, new features which need to be created because of the systems approach or local school context. Co-adaptation and co-design proposals will be finalised in consultation with our youth and advisory boards.

##### Co-adaptation and co-design

Our aim in this stage is to optimise the acceptability, feasibility, efficacy and meaningful evaluation of the suite of interventions by involving end-users and stakeholders in intervention adaptation and its associated implementation approach. We will host co-design / co-adaptation workshops for each WP intervention, each involving adolescents (14–16y, single gendered groups with more females), parents, schools (including teachers and institute heads) and key stakeholders in separate workshops. WP2 co-design will additionally involve national organisations (e.g. State Education & Health Department, Teachers of India, ChildLine India, and Save the Children). WP4 co-design will begin with a stakeholder event to identify community-led solutions to reaching parents. Creative evaluation methods will be devised during co-design to be sensitive to low literacy levels, low chance of measure completion and desirability bias. Implementation barriers, facilitators and developing implementation protocol will be a standing item for consultation in co-design workshops. Co-design / co-adaptation workshops will include a range of age- and role-appropriate discussions and activities centering on the co-adaptation proposals developed in the previous stage.

#### Phase 3: Undertaking Modifications

We will modify the interventions based on co-adaptation and co-design workshop outputs. We will utilise the expanded FRAME approach to report modifications to interventions. [47] This approach documents when, why and how modifications were made, and differentiates cultural adaptation from adaptations made for other reasons. Proposed modifications will be reviewed by the full team to ensure modified interventions still meet the multi-level needs assessment, retain active ingredients and are coherent as a suite of interventions.

Modifications and final prototype interventions will be reviewed by our youth and advisory boards as well as a board of in-country mental health professionals and education sector representatives to ensure they meet state and national requirements. Once finalised, we will produce for each WP intervention a logic model, programme manual, intervention resources, lay counsellor training and supervision protocol, implementation and evaluation protocols.

### Year 2 Feasibility testing

Lay counsellors will receive full training prior to the launch of the feasibility study. The feasibility study will run in eight schools, with three randomly assigned to waitlist. The four interventions will run in parallel in schools. Lay counsellors will complete weekly monitoring logs of intervention delivery in each school, including logging of modifications made (as per FRAME [47] and will have weekly supervision by clinical team members. Fidelity monitoring will be undertaken by a team of trained data collection assistants.

We have established preliminary feasibility indicators for each intervention, which will be updated when interventions are finalised. Feasibility indicators which will be common across WP1–4 interventions are: meeting recruitment and retention rates, delivery as intended, measure completion and no significant adverse effects. We will adopt a ‘traffic-light’ system for recruitment and retention as follows: Red – if rates below 50% (say) then will not proceed; Amber – if rates 50-65% then proceed if there are plans to improve; Green – above 65% proceed to definitive trial. Mixed-methods process and evaluation data will be secured for all interventions from adolescents, teachers, parents and lay counsellors to explore: perceived usefulness of the interventions; implementation challenges; unintended negative effects; safeguarding challenges; the process of change and intervention development needs. Evaluation outcomes will inform intervention improvements and parameters for a definitive trial.

#### Sample size calculation

The planned sample for the feasibility study by intervention is detailed in Table 2. Calculation of power is based on WP1 and on an analysis of variance approach for simplicity. We will test two candidate primary outcomes measures for WP1, the Strengths and Difficulties Questionnaire (SDQ) [48] and the Revised Child Anxiety and Depression Scale (RACDS) [49]. SDQ is an emotional and behavioural screening tool, culturally validated in India and used widely in adolescent mental health research. RCADS measures anxiety and depression and demonstrates excellent sensitivity-to-change. A translated RCADS has been used with Indian populations [50] but full cultural validity needs to be established. As this will be an important tool to strengthen Indian mental health research, we will conduct a full validation study in parallel to the co-design stage. We have determined via a UK school feasibility trial [51] that the change in SDQ had a standard deviation of 2.8 points. Assuming 80% recruitment, comparing the mean changes in SDQ from 480 participants from the intervention group with 288 from the waitlist group using a 1% significance level has more than 98% power to detect a difference of 1 point on the SDQ scale. Around 960 adolescents will be approached for WP1. At 65% recruitment, the power remains over 95%.

**Table 2:** Planned sample size for each intervention in the feasibility study.

| WP | Intervention | N | Sample |
| --- | --- | --- | --- |
| 1 | Universal curriculum adolescent mental health programme | 960 | Grade 9 15–16-year-olds from 8 schools (120 per school) |
| 2 | Teacher education programme | 32 | Grade 9 teachers (4 per school) |
| 3 | School climate intervention | 8 schools | To be defined following intervention selection |
| 4 | Parental mental literacy intervention | 960 sets of parents | Aim to reach all parents / carers of WP1 participants |

#### WP5 Implementation research

We will test the feasibility of our co-designed implementation approach, which will draw on improvement science. Lay counsellors will be trained and supported to optimise the intervention in situ by logging and overcoming implementation challenges as they arise, whilst keeping fidelity to intervention function. Process evaluations based on interviews / focus groups and implementation assessment tools will be conducted post-intervention with key school staff and other stakeholder as needed depending on what challenges arise. Solutions to implementation challenges will be sought.

#### WP6 Health economic evaluation

Using a micro (bottom-up) costing strategy [52], separate design and delivery costs per intervention component will be estimated using available data sources (e.g. government ministry data, national pay scales, national earnings surveys) to determine unit costs for personnel time. Any additional primary data needs will be identified. Costs will be presented at the class and school level to promote generalisability. Preference-based outcome health and wellbeing measures are needed for economic evaluation [53] but there is limited evidence on their use with adolescents and no evidence of their use feasibility in this context (with adolescents in school-based settings and in India). We will feasibility test candidate measures including CHU-9D [54] and EQ-5D-Y [45] via pre-post administration to 50 intervention adolescents and via 10 think-aloud interviews to identify completion challenges. We will also administer the EQ-5D-5L [56] and ICECAP-A [57] pre-and post-intervention to 50 parents to determine measure completion and any need for instrument adaptation. We will use the cost and outcome data to generate exploratory cost-consequence analyses of the intervention strategies.

#### WP7 Understanding barriers and opportunities for research to policy uptake

We aim to (i) understand facilitators, barriers and solutions to policy uptake of the evidence on school mental health (in Karnataka); (ii) identify ways of increasing adolescents’ participation in policy processes, and (ii) identify the potential for application of solutions to other states. A policy and stakeholder mapping exercise will be undertaken, and then key advisors (n∼10) for each of the three policies (two National and one state) [29] will be purposefully identified and interviewed. Insights will be shared at a stakeholder event comprising about 20 education and health decision-makers, adolescents and teachers to generate solutions for policy improvement. Insights will direct a social media campaign for policy action. Evaluation of effectiveness will be based on the number and nature of stakeholders reached, new knowledge on evidence-to-policy barriers and solutions and adolescents’ participation.

#### WP8 Community Film making and social media campaign

Around 15 adolescents (single-gendered groups and more females) will be supported to create 3 short films on experiences and risks around adolescent mental health, the potential for the school to affect mental health, and solutions to safeguard their well-being. Films will be used in WP1-4 for engagement, education, advocacy and impact, and to inform (i) a community and stakeholder’s event. Films will be used in WP1-4 for engagement, education, advocacy and impact, via (i) a community and stakeholder’s event to screen the films to promote shared understanding and solutions; (ii) a social media education campaign targeting adolescents’ mental health literacy; and (iii) an advocacy campaign targeting national-level policymakers. Outcomes will include open access films and new knowledge on the feasibility and reach of community filmmaking for health education and advocacy campaigning for school mental health in India.

### Data analysis plan

#### WP1-5 Co-design / co-adaptation

Workshop data will include audio / video recordings, facilitator notes and design materials (e.g. maps, user journeys, reference ranking, annotated implementation and safeguarding protocols). Key workshop moments which informed an adaptation or new design component will be transcribed, supplemented with facilitator notes and accuracy checked by two facilitators. Workshop data from different stakeholder groups per focus (e.g. intervention, implementation, safeguarding) will be integrated by WP leads. They will synthesise proposed modifications and report them as per FRAME [47] to take forward to final approval stages in preparation for the feasibility study (described above).

#### WP1–5 Feasibility Study

School-level effects will be handled as fixed rather than random effects. Primary analysis will be analysis of covariance where outcomes at three months are regressed upon the baseline value and any relevant covariates/factors as age, sex, school, participant characteristics. The analysis will be supported by a two-level mixed-effects model where measures at two-time points are nested within participants. If further to this analysis, a model is required to examine the relationship between the change in measure over time and the initial measure level, then Bloomqvist’s method will be employed. Process evaluations will be analysed using Framework Analysis [58] or Thematic Analysis [59]. The implementation achievements in each school will be quantified per intervention and as a systems approach, and actions associated with success will be identified, drawing on the CFIR [44] and as per Hudson et al. [43].

#### WP6 Health economic evaluation

For the quantitative validity testing of the outcomes measures, we will calculate completion rates of the measures and items. For the qualitative validity testing. we will code the think-aloud interview transcripts to identify errors or difficulties in responding to items. Mean pre- and post-quality of life scores will be used to estimate quality of life benefits from the interventions. These data will be combined with estimates of the intervention costs in a simple cost–consequence analysis [60].

#### WP7 Understanding barriers and opportunities for research to policy uptake

Data from the stakeholder analysis and document reviews will be continuously triangulated with analysis of transcripts from the in-depth interviews. All qualitative data will be analysed using Framework Approach [58] and guided by the three questions for WP7 and relevant theoretical frameworks for evidence-informed policymaking. [61-64]

### Patient and public involvement

Throughout SAMA, listening to adolescents will be central and incorporated in each WP. There are many barriers to adolescent involvement in low resource settings.[31] These can include high levels of mental health stigma as well as cultural differences where adolescent’s opinions might be less commonly sought and heard. Our approach to adolescent participation will be informed by the WHO Global Consensus Statement on Meaningful Adolescent & Youth Engagement. [65] Adolescents will be trained and supported to contribute to the project. As well as being involved as participants in the design and evaluation of interventions in WP1-5, other adolescents will sit our Youth Advisory Panel to act as ‘critical friends’ of the project. Adolescent contribution will also be via participatory filmmaking. This is a key platform within and beyond our project for youth voice in India on adolescent mental health and the role of schools in wellbeing. Other stakeholders including parents, teachers, the school community and policymakers will be involved in the co-design and evaluation stages of the study.

## Data Availability

Certain data will be available on a data sharing site at study end.

## Ethics and dissemination

This research is being conducted in community settings and has received ethical approval from the National Institute of Mental Health Neurosciences Research Ethics Committee (NIMHANS/26^th^ IEC (Behv Sc Div / 2020/2021)), and the University of Leeds School of Psychology Research Ethics Committee (PSYC-221). Additional approvals have been granted by the Karnataka state primary and secondary education ministry and the Karnataka Department of State Education Research and Training.

We will seek consent from parents, teachers, school staff and other stakeholders and assent from adolescents. Grade 9 pupils will receive WP1 and 3 but can opt out of data collection. This is to ensure that participants’ rights are not undermined by universal delivery. Teachers can access WP2 training without consenting to data collection. This is in recognition of the sensitivity of teachers reporting their mental health knowledge or use of harsh discipline practices, and reponses will inform our understanding of the acceptability of our data collection measures. Audio/video recording in workshops and for WP8 will require explicit consent. Research data will be kept secure and confidential, with agreed UK–India data management plans and data sharing agreements.

Alongside WP1–5, we will be drafting, and refining through co-design, a stepped response and referral protocol to be assessed as part of the feasibility study. This protocol will draw on national and international guidance, as well as established practice in NIMHANS. Our priorities are response protocols for identification of (i) moderate to serious mental health need or risk (especially suicide risk) and (ii) abuse (especially sexual abuse). The iterative production and field testing of this school mental health safeguarding policy will be an important contribution to the research field as well as to school systems.

Our plans for dissemination include: (i) events – launch, film screening, impact and capacity building events; (ii) website – project website with multiple functionalities to engage, share resources, disseminate findings and in ways that can be sustained beyond the project; (iii) social media (twitter, Instagram and TikTok) to reach multiple sectors and adolescents; (iv) project briefs/updates/newsletters (flyers), especially for schools and community organisations; (v) executive reports to key stakeholders (e.g. education and health departments, teacher training colleges, mental health service providers); (vi) sharing the project on global repositories, such as the Mental Health Innovation Network; (vii) using existing communication platforms from University of Leeds Global Challenges project relevant to adolescent wellbeing in LMICs; and (viii) conferences and peer-reviewed publications.

## Authors contributions

All authors contributed to the design of the protocol and approved the final version. SHJ and PM produced early manuscript drafts and all other authors reviewed and edited the manuscript.

